# Dynamics of humoral and cellular immune responses after homologous and heterologous SARS-CoV-2 vaccination with ChAdOx1 nCoV-19 and BNT162b2

**DOI:** 10.1101/2022.03.23.22272771

**Authors:** Emanuel Vogel, Katharina Kocher, Alina Priller, Cho-Chin Cheng, Philipp Steininger, Bo-Hung Liao, Nina Körber, Annika Willmann, Pascal Irrgang, Jürgen Held, Carolin Moosmann, Vivianne Schmidt, Stephanie Beileke, Monika Wytopil, Sarah Heringer, Tanja Bauer, Ronja Brockhoff, Samuel Jeske, Hrvoje Mijocevic, Catharina Christa, Jon Salmanton-García, Kathrin Tinnefeld, Christian Bogdan, Sarah Yazici, Percy Knolle, Oliver A. Cornely, Klaus Überla, Ulrike Protzer, Kilian Schober, Matthias Tenbusch

**Author notes:** **Corresponding author(s):** Ulrike Protzer, Kilian Schober & Matthias Tenbusch. These authors contributed equally (Emanuel Vogel, Katharina Kocher, Alina Priller, Cho-Chin Cheng, Ulrike Protzer, Kilian Schober, Matthias Tenbusch).

## Abstract

Vaccines are the most important means to overcome the SARS-CoV-2 pandemic. They induce specific antibody and T-cell responses but it remains open how well vaccine-induced immunity is preserved over time following homologous and heterologous immunization regimens. Here, we compared the dynamics of humoral and cellular immune responses up to 5 months after homologous or heterologous vaccination with either ChAdOx1-nCoV-19 (ChAd) or BNT162b2 (BNT) or both. Antibody responses significantly waned after vaccination, irrespective of the regimen. The capacity to neutralize SARS-CoV-2 – including variants of concern such as Delta or Omicron – was superior after heterologous compared to homologous BNT vaccination, both of which resulted in longer-lasting humoral immunity than homologous ChAd immunization. T-cell responses showed less waning irrespective of the vaccination regimen. These findings demonstrate that heterologous vaccination with ChAd and BNT is a potent approach to induce long-term humoral and cellular immune protection.

**Research in context:** *Evidence before this study:* Due to some rare severe side effects after the administration of the adenoviral vaccine, ChAdOx1 nCoV-19, many countries recommended a heterologous vaccination scheme including mRNA vaccines like BNT162b2 for the second dose. We performed a PubMed search (with no restrictions on time span) using the search terms “SARS-CoV-2” and “heterologous vaccination” and obtained 247 results. Only a fraction of manuscripts included direct comparisons of patient cohorts that received either a heterologous or a homologous vaccination regimen. Of those, the vast majority investigated only short-term immunogenicity after vaccination. Thus, little is known about the long-term maintenance of immunity by heterologous compared to homologous vaccination.

*Added value of this study:* We add a very comprehensive and comparative study investigating heterologous and homologous vaccination regimens early and late after vaccination. Key features include the number of patients (n = 473), the number of vaccination cohorts (n= 3), the fact that samples were derived from three independent study centers and comparative analyses were performed at two independent study centers, as well as in-depth investigation of humoral and T cellular immunity.

*Implications of all the available evidence:* The recent data creates a line of evidence that heterologous vaccination, compared to homologous vaccination regimens, results in at least non-inferior maintenance of humoral and cellular immunity. The enhanced understanding of immunity induced by individual vaccination regimens is crucial for further recommendations regarding the necessity, timing and choice of additional vaccinations and public health policies.

## Introduction

The widespread use of safe and effective vaccines is essential for overcoming the Severe Acute Respiratory Syndrome Coronavirus 2 (SARS-CoV-2) pandemic. As of today, billions of doses of Coronavirus Disease 19 (COVID-19) vaccines, based on adenoviral vectors or mRNA, have been administered worldwide. In very rare cases, the administration of the adenoviral vector-based ChAdOx1-nCov-19 (ChAd) vaccine has been associated with the induction of a vaccine-induced thrombocytopenic thrombosis syndrome, particularly in young women^1^. Consequently, the vaccination authorities of several countries recommended that persons under the age of 60 years who had received a primary dose of ChAd should receive an mRNA-based Covid-19 vaccine for the second immunization^2^.

We and others have previously shown that the heterologous combination of ChAd and mRNA vaccination results in a non-inferior or even superior humoral and cellular immune response compared to homologous mRNA or ChAd vaccination regimens^3–12^. While homologous ChAd vaccination elicited a strong T-cell response shortly after the second immunization, antibody responses were inferior to homologous or heterologous regimens with mRNA vaccines. Furthermore, in the case of homologous vaccination regimens, various studies have shown a decline in antibody and T-cell levels a few months after the second dose^13^. For heterologous vaccination regimens, however, long-term follow-up data on how long B- and T-cell immunity persists are limited^14,15^. This particularly applies to the immune response against newly emerged SARS-CoV-2 variants of concern (VoC) such as the Delta or Omicron mutant^8^. Currently, it therefore remains unclear how the heterologous combination of ChAd and mRNA vaccination compares to homologous mRNA or ChAd vaccination in terms of long-term maintenance of humoral and cellular immunity.

Here, we examined humoral and cellular immunity in up to 473 participants from three different study centers at different time points before, and up to 5 months after heterologous and homologous vaccination with mRNA with BNT162b2 (BNT) and ChAd. While T-cell responses showed only modest contraction, significant waning of humoral immunity was observed over time in all three vaccination regimens. Compared to homologous vaccination with ChAd or BNT, the heterologous regimen generally resulted in more constant antibody responses both in terms of quantity and quality.

## Results

### Heterologous COVID-19 vaccination induced strong antibody responses which are superior or comparable to homologous mRNA vaccination regimens

We compared humoral and cellular immune responses in 473 healthy individuals about 2 weeks (“early after #2”) and 3.5 to 5 months (“late after #2”) after heterologous or homologous vaccination with ChAd and BNT (Table 1). Previously, we had reported limited data on antibody responses early after second vaccination^5^. In the present analysis, median time points for the “early after #2” and “late after #2” analyses were 13-15 and 98-158 days after second vaccination, respectively. Analyses were performed at the study centers in Munich (Munich and Cologne samples) and Erlangen (Erlangen samples) (Table 1). The results are presented by study center for better comparability.

**Table 1:**
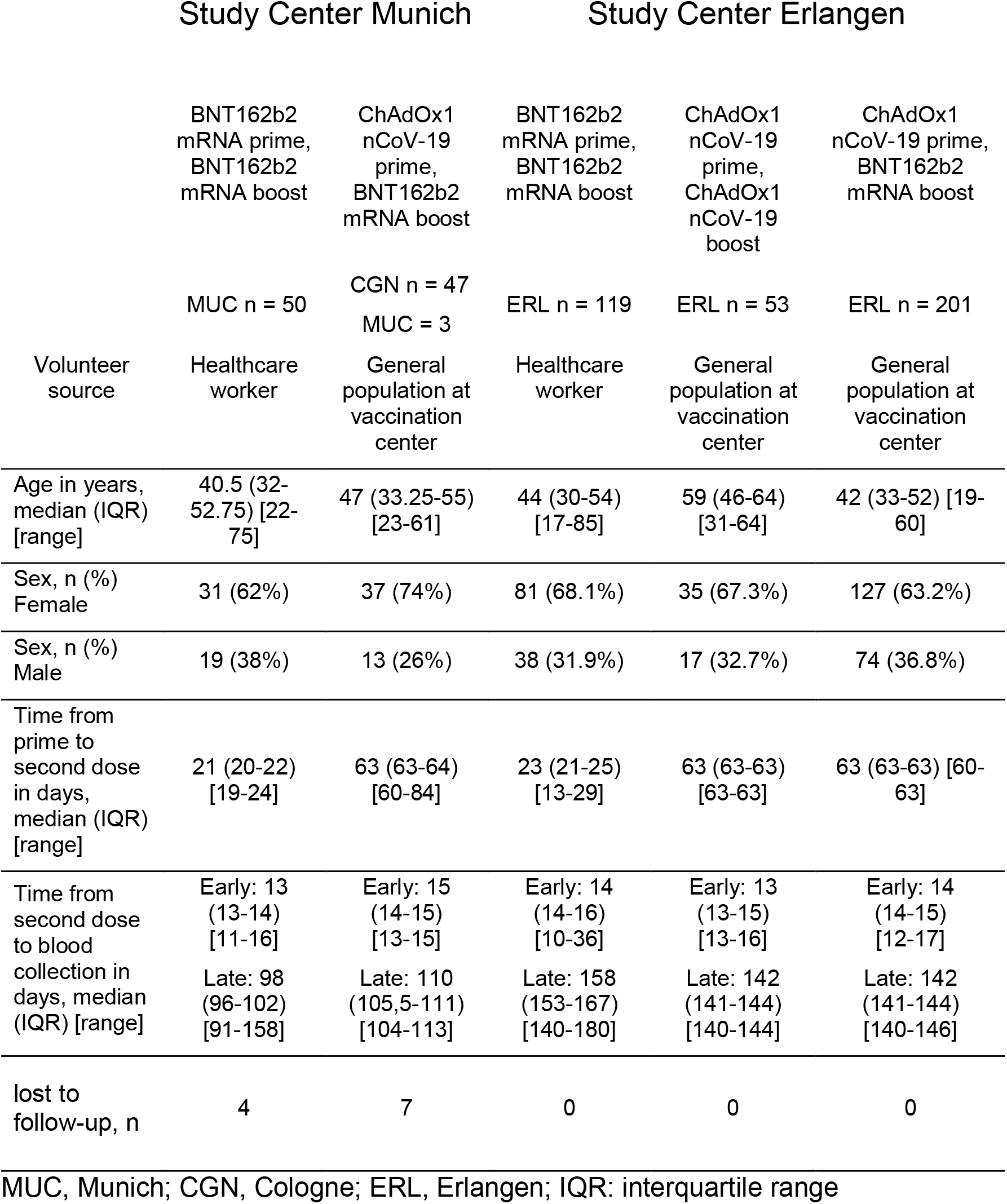
Detailed representation of different study cohorts, separated by study center.

|  | Study Center Munich |  | Study Center Erlangen |  |  |
| --- | --- | --- | --- | --- | --- |
|  | BNT162b2 mRNA prime, BNT162b2 mRNA boost | ChAdOx1 nCoV-19 prime, BNT162b2 mRNA boost | BNT162b2 mRNA prime, BNT162b2 mRNA boost | ChAdOx1 nCoV-19 prime, ChAdOx1 nCoV-19 boost | ChAdOx1 nCoV-19 prime, BNT162b2 mRNA boost |
|  | MUC n = 50 | CGN n = 47<br>MUC = 3 | ERL n = 119 | ERL n = 53 | ERL n = 201 |
| Volunteer source | Healthcare worker | General population at vaccination center | Healthcare worker | General population at vaccination center | General population at vaccination center |
| Age in years, median (IQR) [range] | 40.5 (32-52.75) [22-75] | 47 (33.25-55) [23-61] | 44 (30-54) [17-85] | 59 (46-64) [31-64] | 42 (33-52) [19-60] |
| Sex, n (%)<br>Female | 31 (62%) | 37 (74%) | 81 (68.1%) | 35 (67.3%) | 127 (63.2%) |
| Sex, n (%)<br>Male | 19 (38%) | 13 (26%) | 38 (31.9%) | 17 (32.7%) | 74 (36.8%) |
| Time from prime to second dose in days, median (IQR) [range] | 21 (20-22) [19-24] | 63 (63-64) [60-84] | 23 (21-25) [13-29] | 63 (63-63) [63-63] | 63 (63-63) [60-63] |
| Time from second dose to blood collection in days, median (IQR) [range] | Early: 13 (13-14) [11-16]<br>Late: 98 (96-102) [91-158] | Early: 15 (14-15) [13-15]<br>Late: 110 (105,5-111) [104-113] | Early: 14 (14-16) [10-36]<br>Late: 158 (153-167) [140-180] | Early: 13 (13-15) [13-16]<br>Late: 142 (141-144) [140-144] | Early: 14 (14-15) [12-17]<br>Late: 142 (141-144) [140-146] |
| lost to follow-up, n | 4 | 7 | 0 | 0 | 0 |

MUC, Munich; CGN, Cologne; ERL, Erlangen; IQR: interquartile range

We first assessed the quantities of antibody levels by a surrogate neutralization assay (sVNT) (Fig. 1). This assay correlates well with a real virus neutralization assay not only for the EU-strain SARS-CoV-2 D614G virus, as previously described^5^, but also for the Delta VoC (Suppl. Fig. 1). Regardless of the vaccination schedule, we observed significant waning of antibody levels in almost all individuals at the late time point (Table 1).

**Figure 1:**
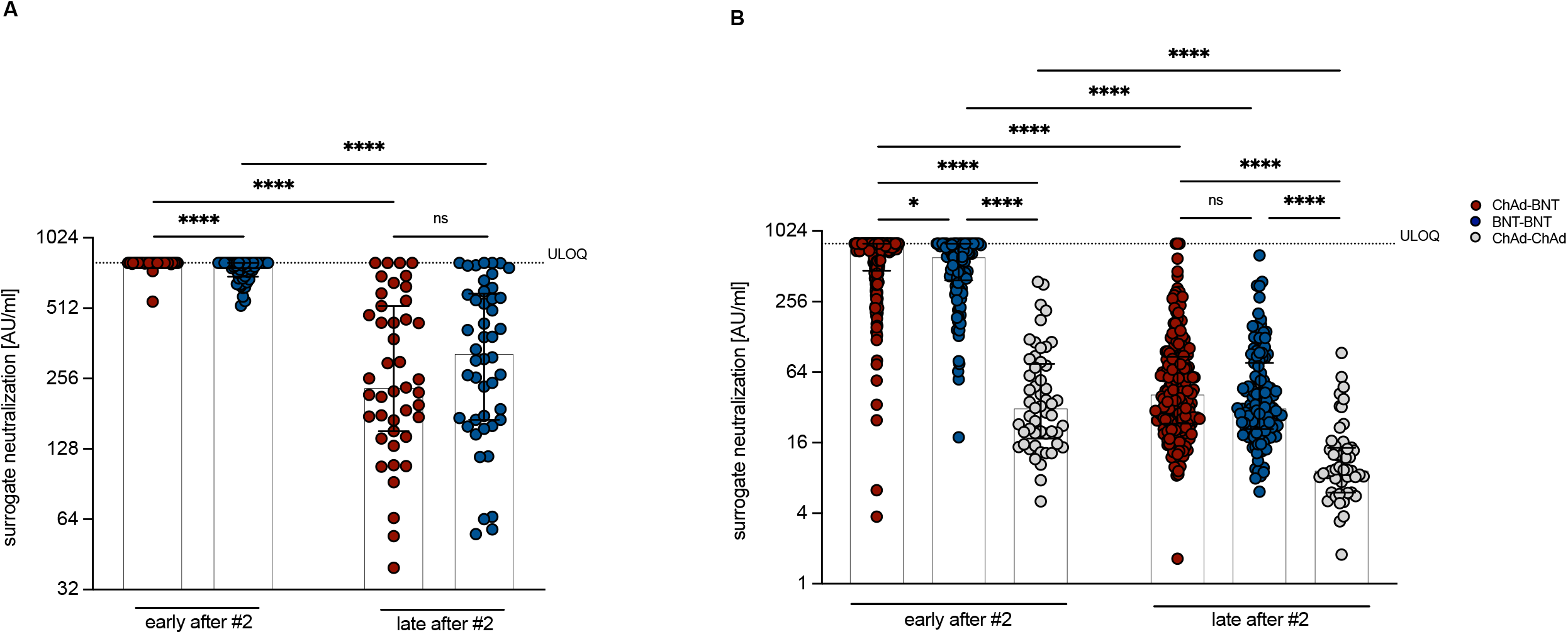
Quantitative antibody levels after heterologous ChAd-BNT vaccination are non-inferior compared to homologous vaccination regimens. Surrogate virus neutralization levels measured at the Munich (A) and Erlangen (B) study site after heterologous ChAd-BNT or homologous BNT-BNT or ChAd-ChAd vaccination. The sVNT is only validated up to a maximum of 800 AU/ml, therefore all values measured as greater than 800 AU/ml were set to 800 AU/ml. (A) „Early after #2” refers to a median of 15 days (for BNT-BNT) or 13 days (for ChAd-BNT) after the second vaccination. „Late after #2” refers to sampling at median 98 days (for BNT-BNT) or 110 days (for ChAd-BNT) after the second vaccination. n = 50 and 43 (ChAd-BNT; “early after #2” and „late “, respectively) and n = 50 and 46 (BNT-BNT; “early after #2” and „late after #2”, respectively). (B) „Early after #2” refers to sampling at median 14 days (for BNT-BNT and ChAd-BNT) and 13 days (for ChAd-ChAd) after the second vaccination. „Late after #2” refers to a median of 158 days (for BNT-BNT) or 142 days (for ChAd-BNT and ChAd-ChAd) after the second vaccination. n = 201 (ChAd-BNT), 119 (BNT-BNT) and 53 (ChAd-ChAd). ULOQ = upper limit of quantification (800 AU/ml). For inter-group statistics concerning one time point Mann-Whitney (A) or Kruskal-Wallis followed by Dunn’s multiple comparisons test was used (B). Bars represent group medians, whiskers interquartile range. Over-time comparison within one group was done by Wilcoxon test. *p < 0.05, **p < 0.01, ***p < 0.001, ****p < 0.0001, and n.s. indicates not significant. A detailed description of the data can be found in Supplemental Table for Figure 1A and B.

At the study center in Munich (Fig. 1A), antibody neutralization capacity at the follow-up time point was significantly reduced compared to the time point early after second vaccination, but the remaining antibody levels were similar after ChAd-BNT (median = 234.65 AU/ml; n = 43) and BNT-BNT (median = 328.17 AU/ml; n = 46) vaccination. At an independent study center in Erlangen, these results were confirmed (Fig. 1B). Furthermore, additional analysis of samples from a homologous ChAd-ChAd vaccination scheme cohort showed that neutralizing antibody levels late after homologous ChAd-ChAd vaccination (median of 9.32 AU/ml; n = 53) were still significantly lower compared to homologous BNT-BNT (median = 31.74 AU/ml; n = 119) or heterologous ChAd-BNT (median = 41.72 AU/ml; n = 201) vaccination (Fig. 1B). Thus, the heterologous ChAd-BNT vaccination regimen results in long-term neutralizing antibody levels against SARS-CoV-2 WT virus which are as high as after homologous BNT-BNT and higher than after homologous ChAd-ChAd immunization.

### Serum neutralization capacity of variants of concern is superior after heterologous vaccination

To investigate potential differences in serological responses between the different cohorts in more detail, we next applied a “real” virus infection neutralization assay (rVNT) for the most relevant SARS-CoV-2 VoC. At the Munich study center, neutralization of VoC B.1.617.2 (Delta) and B.1.1.529 (Omicron) was investigated using sera collected 2 weeks and 3.5 months after heterologous ChAd-BNT or homologous BNT-BNT vaccination (Fig. 2A). Early after the second vaccine dose, heterologous ChAd-BNT vaccination resulted in significantly better serum neutralization capacity of Delta, and to a lesser extent also Omicron, than homologous BNT-BNT vaccination (ChAd-BNT median IC_50_ = 929.15; n=50; BNT-BNT median IC_50_= 432.85; n=50). Serum neutralization capacity for Omicron compared to Delta was reduced 25.8-fold and 21.6-fold for ChAd-BNT (median IC_50_ = 36) and BNT-BNT (median IC_50_ = 20), respectively, in an analysis of sub-cohorts consisting of 15 participants each. 3.5 months after the second vaccination, serum neutralization capacity for Delta still significantly differed between ChAd-BNT (median IC_50_= 370.45, n = 43) and BNT-BNT vaccination (median IC_50_ = 72.92, n = 46). However, there was barely any neutralization capacity left against Omicron in either cohort (Fig. 2A).

**Figure 2:**
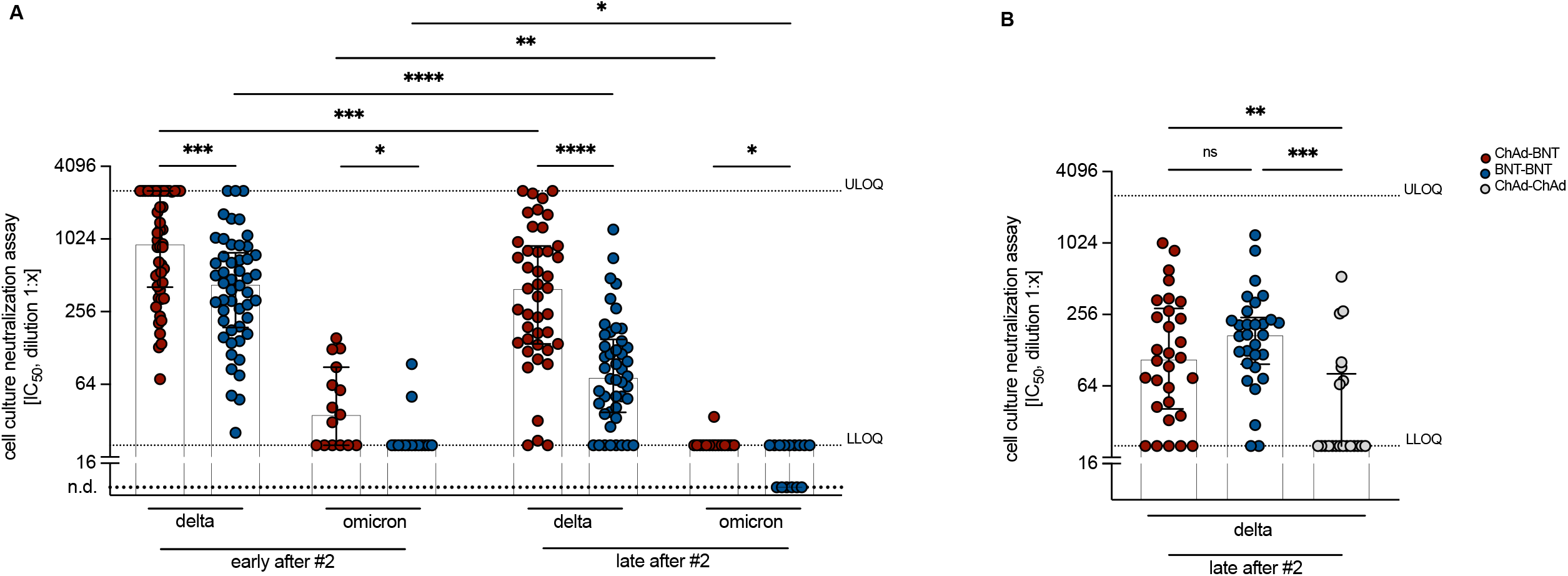
Individuals of the heterologous cohort neutralize variants of concerns more efficiently than individuals from homologous cohorts. Real virus neutralization levels measured against Delta and Omicron at the Munich (A) or Erlangen (B) study site after heterologous ChAd-BNT or homologous BNT-BNT or ChAd-ChAd vaccination. (A) „Early after #2” refers to on average (median) 13-15 days after second BNT vaccination. „Late after #2” refers to on average (median) 98-110 days after second vaccination. n = 50 and 43 (ChAd-BNT; “early after #2” and „late after #2”, respectively) and n = 50 and 46 (BNT-BNT; “early after #2” and „late after #2”, respectively). Each group was measured against Delta and Omicron at each time point. (B) „late after #2” refers to on average (median) 158 days after second BNT (for BNT-BNT) and 142 days after second BNT or ChAd (for ChAd-BNT and ChAd-ChAd) vaccination. n = 30 (ChAd-BNT), n = 30 (BNT-BNT) and n = 21 (ChAd-ChAd). Here, only neutralization of Delta was tested. Bars represent group medians, whiskers interquartile range. ULOQ = upper limit of quantification (2560). LLOQ = lower limit of quantification (20). n.d. = not detected. (A) For inter-group statistics Kruskal-Wallis followed by Dunn’s multiple comparisons test was used (B). Over-time comparison within one group was done by Wilcoxon test ((A) (B)). *p < 0.05, **p < 0.01, ***p < 0.001, ****p < 0.0001, and n.s. indicates not significant. A detailed description of the data can be found in Supplemental Table for Figure 2A and B.

To confirm these results, serum samples from the Erlangen study center collected at 4.5 to 5 months after second vaccination („late after #2”) were analyzed in the rVNT assay for the ability to neutralize the Delta variant. These analyses again additionally included a cohort of homologous ChAd-ChAd vaccinated participants. Late after ChAd-ChAd immunization, barely any neutralization capacity against Delta was detectable (median IC_50_ = 20; n = 21), which was significantly different from the ChAd-BNT and BNT-BNT cohorts. In contrast to the results obtained at the Munich study center, there was no significant difference in neutralization capacity against Delta between the ChAd-BNT (median IC_50_ = 107.8; n = 30) and BNT-BNT (median IC_50_ = 172; n = 30) group (Fig. 2B). These findings were further confirmed using a pseudovirus neutralization assay (pVNT), although overall the neutralization titers were slightly lower than in the rVNT (Suppl. Fig. 2). Whether this is due to the later sampling time point, the lower numbers of participants, or another reason, cannot be clarified. The reduced capacity to recognize the spike (S) proteins of VoC in comparison to the S protein of the original Wuhan strain observed in the pVNT assay (Suppl. Fig. 2) was confirmed by a flow cytometric analysis using HEK293 cells expressing the corresponding S proteins in their natural conformation on the cell surface (Suppl. Figure 3). Overall, these results demonstrate that humoral immunity against VoC was reduced irrespective of the vaccination regimen, but that differences in antibody neutralization capacity between the immunization cohorts remained unchanged.

### Heterologous vaccination results in increased antibody avidity

The neutralization capacity of antibodies depends not only on their quantity, but also on their quality^16^. We therefore next applied a modified quantitative anti-S ELISA to determine antibody avidity against the S1 domain of the SARS-CoV-2 WT spike antigen. To this end, we used samples of the sub-cohorts from the Omicron rVNT analysis (Fig. 3). 2 weeks after second vaccination we observed a higher avidity of antibodies in the heterologous ChAd-BNT (median = 57.96 %) compared to the homologous BNT-BNT (median = 30.86 %) cohort. This difference remained constant at follow-up (Fig. 3). Over time, there was a tentative increase in antibody avidity for both groups (BNT-BNT median = 49.49 %; ChAd-BNT median = 65.69 %) which was, however, not statistically significant (Fig. 3). These results indicate that higher antibody avidity after heterologous ChAd-BNT compared to homologous BNT-BNT vaccination contributes to non-inferior neutralization capacity (Fig. 1).

**Figure 3:**
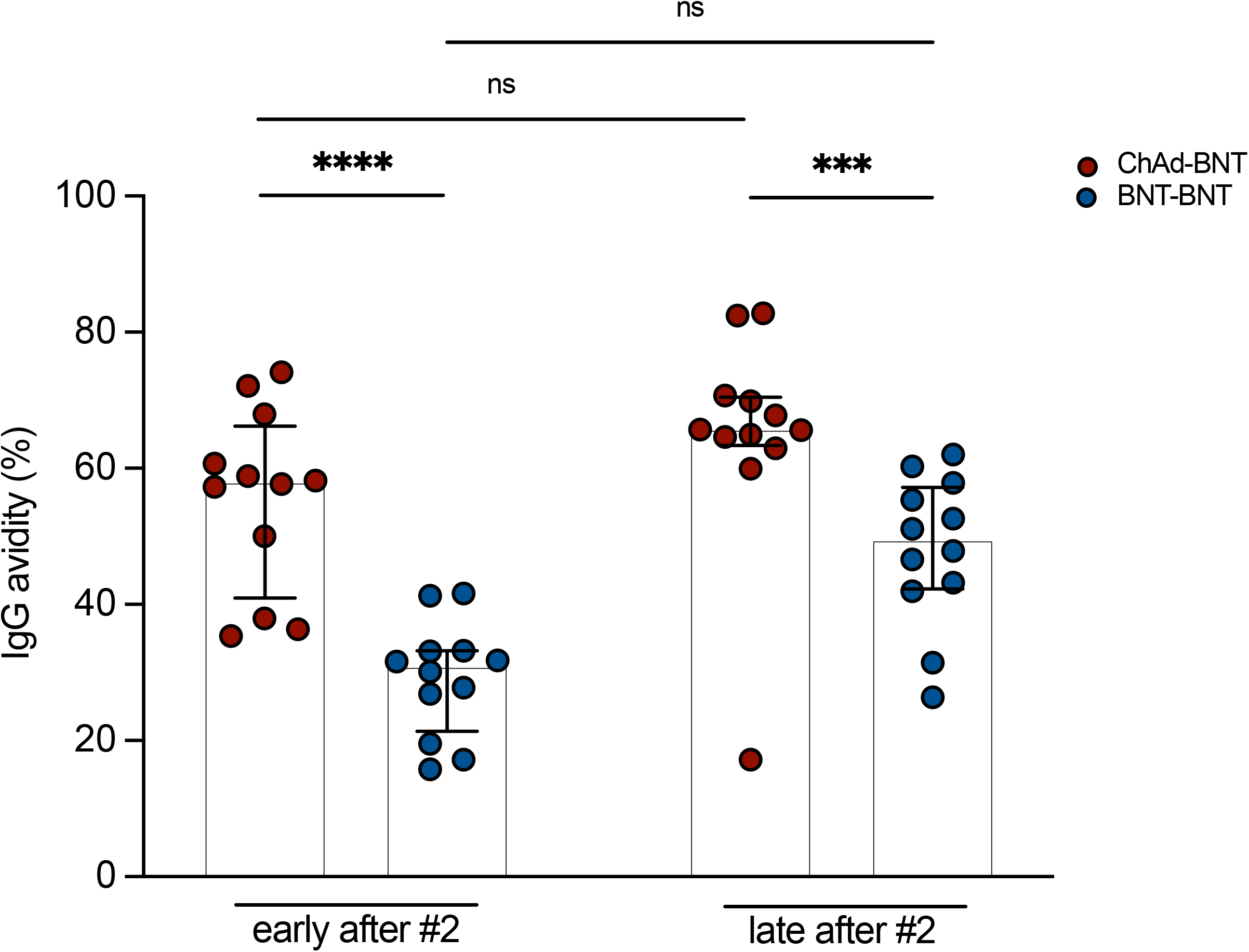
Higher antibody avidity upon heterologous ChAd-BNT compared to homologous BNT-BNT vaccination. Antibody avidity of a subcohort (n = 12) from study center Munich after heterologous ChAd-BNT and homologous BNT-BNT vaccination. „Early after #2” refers to on average (median) 13-15 days after secondary BNT vaccination. „Late after #2” refers to on average (median) 98 and 110 days after second vaccination. Bars represent group medians, whiskers interquartile range. For inter-group statistics concerning one time point Mann-Whitney test was used. Over-time comparison within one group was done by Friedman test. A detailed description of the data can be found in Supplemental Table for Figure 3.

### Homologous and heterologous vaccination induce stable polyfunctional SARS-CoV-2 spike-reactive T-cell responses

Given the critical role of T lymphocytes in protection against SARS-CoV-2 infection, we next also characterized the T-cell response elicited by heterologous or homologous vaccination regimens. We acquired peripheral blood mononuclear cells (PBMCs) from vaccinated individuals at the two study centers and characterized CD4 and CD8 T-cell responses by IFN-γ enzyme-linked immunospot (ELISPOT), IFN-γ/IL-2 Fluorospot as well as intracellular cytokine staining followed by flow cytometry analysis (ICCS). To this end, PBMCs were stimulated overnight with two 15mer peptide pools covering the S1 and S2 domains of the full-length SARS-CoV-2 spike glycoprotein, respectively.

To evaluate the dynamics of spike-specific T cells, the frequency of antigen-reactive, IFN-γ-producing T cells was first longitudinally characterized within the ChAd-BNT in Munich (Fig. 4A) and the BNT-BNT cohort in Erlangen (Fig. 4B) early after first and second vaccination, as well as at the late follow-up time point. Limited T-cell responses to spike peptide stimulation were observed in some individuals already before vaccination (Fig. 4B), which might result from cross-reactive clonotypes derived from exposure to common cold coronaviruses^17,18^. Induction of spike-specific T cells was observed 55-137 days after one vaccination with ChAd (Munich study site, Fig. 4A) or 10 days after one vaccination with BNT (Erlangen study site, Fig. 4B) in almost all individuals (“early after #1”). T-cell responses peaked 12-36 days (Munich study site, Fig. 4A) or 10 days (Erlangen study site, Fig. 4B) after second immunization with BNT (“early after #2”). 4 months after second immunization with BNT (“late after #2”), comparable responses of spike-reactive T cells were observed for both homologous vaccination regimens as well as the heterologous vaccine cohort.

**Figure 4:**
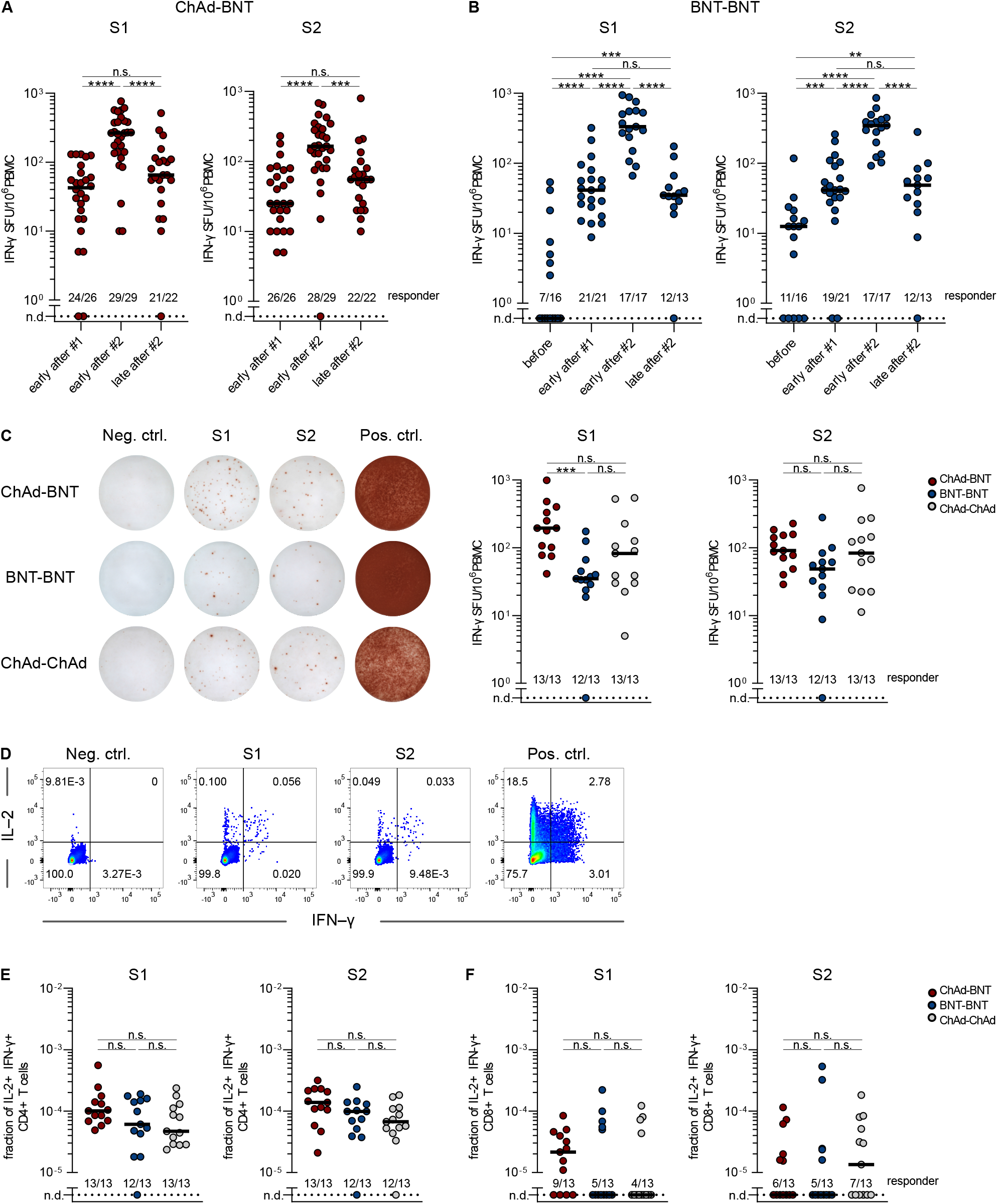
Long-term maintenance of SARS-CoV-2 spike-reactive T cells after homologous and heterologous vaccination. (A)-(B) Longitudinal characterization of spike-specific T cells, quantified by IFN-γ spot forming units (SFU) after stimulation with SARS-CoV-2 spike peptide pools S1 and S2. (A) Vaccinees of Munich study center quantified by IFN-γ Fluorospot. “early after #1”: 55 to 137 days after initial ChAd vaccination, n=26. “early after #2”: 12 to 36 days after second BNT vaccination, n=29. “late after #2”: 91 to 153 days after second BNT vaccination, n=22. (B) Vaccinees of Erlangen study center quantified by IFN-γ ELISPOT. “before”: pre-vaccination, n=16. “early after #1”: 10 days after first BNT vaccination, n=21. “early after #2”: 10 days after second BNT vaccination, n=17. “late after 2#”: 4 months after second BNT vaccination, n=13. The time points “before”, “early after #1”, and “early after #2” refer to a separate cohort of vaccinees that was included for contextualization. (C) Cohort comparison of spike-specific T cells after stimulation with SARS-CoV-2 spike peptide pools S1 and S2, in dilution of solvent (Neg. ctrl.), or with PMA/ionomycin (Pos. ctrl.). Vaccinees of Erlangen study center 4 months after second vaccination (late after #2). Representative data (left) and quantification of IFN-γ SFU for all donors of indicated vaccination cohorts (right) are displayed. (D)-(F) Flow cytometric analyses of polyfunctional spike-specific T cells after stimulation with SARS-CoV-2 spike peptide pools S1 and S2, in dilution of solvent (Neg. ctrl.), or with PMA/ionomycin (Pos. ctrl.). Vaccinees of Erlangen study center 4 month after second vaccination (late after #2). (D) Representative flow cytometry data. Shown gates are pre-gated for CD4+ living lymphocytes. Quantification of IL-2 and IFN-γ double-positive CD4 (E) and CD8 (F) T cells for all donors of indicated vaccination cohorts. Dots represent individual vaccinees. Numbers indicate vaccinees with a positive response defined by a detectable T-cell response above background. Non-responsive vaccinees are represented as not detected (n.d.). For inter-group statistics concerning one time point Kruskal-Wallis test was performed followed by Dunn’s multiple comparisons test ((C), (E), (F)). Over-time comparison within one group was done by Mann-Whitney test ((A), (B)). *p < 0.05, **p < 0.01, ***p < 0.001, ****p < 0.0001, and n.s. indicates not significant.

Having demonstrated that spike-reactive T cells were detectable at least 4 months after the second vaccination, we next examined the effect of different vaccination regimens on the quality of the T-cell response in more detail. We therefore quantified IFN-γ secreting, spike-specific T cells 5 to 6 months after vaccination with the different vaccine regimens in study participants at the Erlangen study center (Fig. 4C). IFN-γ ELISPOT detected reactive T cells in almost all individuals of ChAd-BNT, BNT-BNT and ChAd-ChAd vaccination cohorts. After heterologous ChAd-BNT vaccination, S1-reactive T cells were detected at higher frequencies compared to the homologous BNT-BNT vaccination cohort while there was no difference for S2-specific T cells and the other vaccination schemes. Thus, heterologous vaccination was at least as efficient as homologous vaccination in inducing spike-reactive T-cell responses that are stable over time (Fig. 4C).

T-cell polyfunctionality is a hallmark of high-quality immunity and predictive of protective immune responses^19^. To examine whether heterologous and homologous vaccination regimens induce and maintain polyfunctional T lymphocytes equally well, T cells were characterized for simultaneous production of the effector cytokines IL-2 and IFN-γ (Fig. 4D; Suppl. Fig. 4). Quantification of these double-positive T cells revealed a dominant, polyfunctional CD4 T-cell response that persisted in the majority of individuals irrespective of the vaccination regimen used (Fig. 4E). For CD8 T cells, we observed a greater inter-individual variability (Fig. 4F) with 40-60% not reacting to peptide stimulation at all. This effect was most probably due to variable recognition of CD8 epitopes within the 15mer peptides that were used for antigenic stimulation.

Overall, the frequency of polyfunctional T cells quantified by ICS correlated with the frequency of spike-reactive T cells determined by IFN-γ ELISPOT, further validating the findings (Suppl. Fig. 5). Fluorospot assays further confirmed the induction of IL-2 and IFN-γ secreting polyfunctional T cells after primary immunization with ChAd and secondary immunization with BNT, as well as the persistence of a polyfunctional CD4-dominated T-cell response at the level of primary vaccination throughout the entire observation period (Suppl. Figure 6). In summary, all vaccination regimens induce stable and polyfunctional T-cell responses.

## Discussion

We here analyzed the humoral and cellular immune response of 473 participants from three different study sites, 2 weeks and several months after homologous and heterologous ChAd and BNT vaccination. Overall, heterologous vaccination with ChAd followed by BNT induced equal or even superior humoral and cellular immune responses compared to homologous BNT-BNT or ChAd-ChAd vaccination.

We and others had previously reported enhanced neutralization capacity early after heterologous ChAd-BNT vaccination compared to homologous BNT-BNT-vaccination, both of which in turn induced clearly higher neutralizing antibody titers than a homologous ChAd vaccination regimen^3–12^. Apart from significant waning of neutralization capacity towards WT virus at late time points for all regimens, we here observed that differences in humoral immunity towards WT virus between ChAd-BNT and BNT-BNT vaccination vanished, while homologous ChAd-ChAd still showed reduced neutralization titers compared to the other two vaccination schemes.

In line with previous reports^16,20–30^, S-specific antibodies induced by the current vaccines encoding the S protein from the original Wuhan strain have significantly reduced neutralizing activity against the SARS-CoV-2 VoC Delta, and even less activity against Omicron. Of note, we here detect such loss of neutralization for all vaccination schedules. In terms of differences between the immunization regimens, ChAd-BNT and BNT-BNT groups showed higher neutralizing antibody response against VoCs than the ChAd-ChAd group, as observed for WT virus.

The relative binding capacity to the different spike variants might be also indicative for the degree of immune evasion by the different VoCs. Since neutralizing antibody levels directly correlate with the level of protection against infection^31^, vaccine efficacy (VE) against infection with VoCs also decreased dramatically over time^32–34^. Nevertheless, 3 to 6 months after the second vaccination, VE were reported to be comparable for BNT-BNT and ChAd-BNT, but lower for ChAd-ChAd schedules^35,36^.

Antibody quality might be even more important than the mere quantity for potent vaccine responses, as demonstrated by the high avidity of anti-spike antibodies after a third exposition^16^. In our study, antibody avidity increased slightly from the early to the late time point for both the ChAd-BNT and the BNT-BNT cohort, which might indicate ongoing B-cell maturation. It has been reported that this process could last up to 6 months in recipients of homologous mRNA vaccines or convalescent patients, while comparable data on heterologous vaccinations are missing^37–41^. A higher avidity of antibodies induced by heterologous ChAd-BNT vaccination offers an explanation why they show a superior neutralization capacity against VoC at both study centers. One potential reason for differential affinity maturation of memory B cells is the difference in the interval between the first and second vaccine dose, which was on average 63 days for the ChAd-BNT vaccinees and 21-23 days for homologous BNT-BNT vaccinated individuals. Furthermore, the duration of antigen presentation in the germinal centers might be different after viral vector immunization or mRNA vaccination. The presence of vaccine-derived mRNA and spike protein has been shown in lymph node biopsies from mRNA vaccinated individuals up to 8 weeks^41^. Further investigations to elucidate the differences for the current vaccines are highly relevant for the implementation of future vaccine regimens using gene-based vaccines. Although the VE against symptomatic infections wanes over time due to the reduced neutralizing capacity of vaccine-induced antibodies, and despite the fact that Omicron by now dominates SARS-CoV-2 case numbers worldwide, protection from severe disease progression currently still prevails. Apart from boosters of humoral immunity through a third vaccination, a central reason for this is a more long-lasting^42–46^ and conserved^47–52^ T-cell response. In this context, we also addressed the question to which extent SARS-CoV-2 spike protein-specific T cells will persist in response to different vaccination regimens. Maintenance of spike-reactive T cells was observed for the vast majority of individuals after homologous and heterologous immunization at the late time point. Longitudinal characterization of the frequency in individual vaccinees indicated long-lasting quantities of these spike-specific T cells at a level obtained after the first immunization. This observation was made at both independent study centers. Depending on the readout, the heterologous vaccination regimen was consistently non-inferior and sometimes statistically significantly superior to the homologous BNT immunization. It has already been shown that a priming dose of ChAd induces a stronger T-cell response compared to a primary immunization with BNT, which was however no longer the case after a secondary BNT immunization^53^. Nevertheless, this could still indicate that the overall superiority of humoral and cellular immunogenicity through the heterologous vaccination regimen results from a more potent primary immune response. For example, strong CD4 T-cell responses induced by primary ChAd vaccination may also explain why serological antibody responses after heterologous ChAd-BNT vaccination are more prominent than after homologous BNT-BNT vaccination^53^.

Polyfunctionality as a predictor of an effective T-cell immune response was demonstrated for persisting T cells after all vaccination regimens^19^. Especially polyfunctional CD4 T cells were well maintained 4 months after the second vaccination. For CD8 T cells this was less clear, most probably owing to variable recognition of (shorter) CD8 epitopes within the 15mer peptides that were used for antigenic stimulation. Overall, our data show that heterologous vaccination is at least as capable as homologous vaccination regimens in inducing long-term maintenance of polyfunctional spike-specific T cells, which are likely to convey protective immunity. In summary, these data document at least non-inferior humoral and cellular immunogenicity after heterologous ChAd-BNT vaccination compared to the respective homologous regimens. While waning of humoral immunity and reduced neutralization capacity against VoC was detected for all vaccination regimens, T-cell responses were more consistently conserved. An enhanced understanding of humoral and cellular immunity induced by individual vaccination regimens is crucial for further recommendations regarding the necessity, timing, and choice of additional vaccinations and public health policies.

## Methods

### Study design and participants

The study is a follow-up analysis of 473 homologously or heterologously vaccinated participants that were previously only assessed for the production of antibodies using sVNT^5^. Study participants were divided into three different cohorts according to their vaccination regimen. Subjects of the two homologous groups received two doses of BNT or ChAd, respectively. In contrast, subjects of the third group received the heterologous vaccination regimen consisting of ChAd vaccine for the first and BNT for the second dose. Participants’ sera and peripheral blood mononuclear cells (PBMCs) were analyzed at two different study centers in Germany. Blood sampling schedules varied by study center and cohort. Not all tests were performed at every point in time. In general, four different points in time can be distinguished. Time point “before” is the initial time point at the day of the first vaccination. “Early after #1” refers to the moment of the second vaccination, “early after #2” corresponds to approximately 2 weeks after this vaccination and the “late after #2” time point analysis was carried out between 4 and 5 months after the second vaccination, depending on the study center and vaccination regimen. The cohorts of homologous BNT and ChAd vaccinated people mainly includes healthcare workers, whereas the heterologous vaccinated cohort did not comprise a specific professional group. For longitudinal characterizations of the T-cell responses at the Munich study center (heterologous ChAd-BNT cohort), a separate cohort of vaccinees was included for the time points “early after #1”, “early after #2”, and “late after #2”. For longitudinal characterizations of the T-cell responses at the Erlangen study center (homologous BNT cohort), a separate cohort of vaccinees was included for the time points “before”, “early after #1”, and “early after #2” for contextualization. A detailed description of the cohorts can be found in table 1. Ethics approval was granted by the local ethics committees in Erlangen (Az. 340_21B) and Munich (Az. 26/21 and Az. 330/21 S).

### Antibody response using surrogate virus neutralization assay

We used the iFlash-1800 CLIA Analyzer (YHLO Shenzhen, China) for the quantification of the antibody response. For the detection of neutralizing antibodies, we applied the iFlash-2019-nCoV NAb assay according to the manufacturer’s instructions. The test principle is a competitive immunoassay. The iFlash-2019-nCoV NAb assay is only validated up to a level of 800 AU/ml according to the WHO standard. Therefore, all results exceeding this limit have been set to 800 AU/ml.

### Antibody avidity

Binding strength of the SARS-Cov-2 IgG antibodies was determined by adaptation of the commercial IgG agile SARS-CoV-2 ELISA (Virion/Serion, Germany) using ammonium thiocyanate (NH_4_SCN) (Roth, Germany) as previously described^16,54,55^. Serum samples were measured using the IgG agile SARS-CoV-2 ELISA and diluted to 100 U/mL according to the standard curve provided by the manufacturer to exclude an influence of variable antibody concentrations. Thereafter, serum samples were incubated in the plates pre-coated with Wuhan SARS-CoV-2-spike-ectodomain S1, S2 and BD recombinant antigens for 1h at 37°C in a humid chamber. After washing, antigen-antibody complexes were incubated in the presence of 1.0 M ammonium thiocyanate or PBS as control for 10 min at room temperature. After washing to remove antibodies bound with low avidity, the ELISA was completed according to the manufacturer’s instructions. The relative avidity index was calculated as follows: IgG concentrations (NH_4_SCN) / IgG concentrations (PBS) x 100, and is given in percent.

### Real virus neutralization assay

Based on a previously established infection inhibition assay^16^, VeroE6 cells (ATCC, US) were seeded in 10% fetal calf serum Dulbecco’s Modified Eagles medium (Thermo Fisher Scientific, Germany) at 15,000 cells per well one day before incubation. Infection was started using SARS-CoV-2 at a multiplicity of infection (MOI) of 0.03 plaque-forming units (PFU) / cell. To detect virus-neutralization activity, serum samples were serially diluted 1:2 with DMEM starting from 1:20 up to a 1:2560 or 1:5120 dilution, respectively. SARS-CoV-2 (480 PFU/15,000 cells/well) virus was added in a total volume of 50 µL at 37°C. After one hour of preincubation, the inoculum was transferred to the pre-seeded VeroE6 cells for another one-hour incubation at 37°C before the inoculum was replaced by supplemented DMEM. SARS-CoV-2 infection was terminated after 23 hours by adding 4% paraformaldehyde to fix the cells, and infection rate was analyzed by an in-cell ELISA.

After fixation, cells were washed with PBS and permeabilized with 0.5% saponin (Sigma-Aldrich, Germany). Blocking buffer, consisting of 0.1% saponin-10% goat serum (Sigma-Aldrich) in PBS, was added and incubated for one hour on fixed cells to avoid unspecific binding of antibodies. As a primary antibody, the SinoBiological anti-SARS-CoV-2-N T62 antibody (40143-T62) was used. The antibody was diluted with 1% FCS-PBS to 1:1500 ratio and 50µl were added in each well and incubated at room temperature for 2 hours. After washing, the second antibody was added. Goat anti-rabbit IgG2a-HRP antibody (EMD Millipore / order number 12-348) with 1% FCS-PBS was diluted to 1:4000 ratio. 50µl were added and incubated at room temperature for 1-2 hours. After the final washing step 100µl tetramethylbenzidine (TMB) were incubated for 20 min at room temperature. As final step 2M H_2_SO_4_ were added to stop the reaction. The result was quantified using optical detection with a Tecan Infinite 200 reader (TECAN, Switzerland) at 450 nm wavelength. The inhibition curve of each sample was analyzed by statistical analysis software Graph Pad Prism (GraphPad Software, USA), and 50% inhibitory concentration (IC50) was determined using non-linear regression.

### FACS-based analysis of anti-S binding antibodies

A modified version of our previously published serological assay was used, in which HEK 293T cells either stably expressing the spike protein from the original Wuhan strain or transiently expressing the spike protein of B.1.167.2 or B1.1.529, respectively, were used as target cells^56^. To quantify antigen-specific antibodies, 5×10^5^ HEK 293T cells were incubated with serum samples diluted in 100 µl FACS-PBS (PBS with 0.5% BSA and 1 mM sodium azide) for 20 minutes at 4°C to bind to spike protein on the surface. After washing with 200 µl buffer, bound S-specific antibodies were detected with anti-human IgG-AF647 (4°C, 30 min incubation; clone HP6017, Biolegend, Cat #409320). After further washing, samples were measured on an AttuneNxt (ThermoFisher) and analyzed using FlowJo software (Tree Star Inc.). A standard plasma sample with a defined concentration of 1,01mg/ml anti-SARS-CoV-2S IgG was used as reference control. The median fluorescence intensity (MFI) correlates with the level of bound antibodies^56^.

### Pseudotype neutralization assay

Neutralization of the early D614G (WT) and the B1.617.2 variants was assessed with the help of spike-pseudotyped simian immunodeficiency virus particles as described before^57^. To produce pseudotyped reporter particles, HEK293T cells were transfected with the SIV-based self-inactivating vector encoding luciferase (pGAE-LucW), the SIV-based packaging plasmid (pAdSIV3), and the respective spike variant-encoding plasmid as described previously^58^.

For the assessment of pseudotype neutralization, HEK293T-ACE2 cells were seeded at 2×10^4^ cells/well in a 96well flat bottom plate. 24 h later, 60 µl of serial dilutions of the serum samples were incubated with 60 µl lentiviral particles for 1 h at 37°C. HEK293T cells were washed with PBS and the particle-sample mix was added to the cells. 48 h later, medium was discarded, and the cells washed twice with 200 µl PBS. Following 50µl PBS and 25µl ONE-Glo™ (Promega Corp, Madison, USA) was added and after 3 minutes the luciferase signal was assessed on a microplate luminometer (VICTOR X5, PerkinElmer) and analyzed using PerkinElmer 2030 Manager software. The reciprocal serum ID_50_ was determined with Prism GraphPad 9 (San Diego, California, USA) by application of the Sigmoidal 4PL function. For sera that did not reach neutralization by at least 50% at the highest serum dilution, the ID_50_ was set to the highest reciprocal serum dilution, namely 20.

### Isolation and cultivation of peripheral blood mononuclear cell (PBMC)

PBMCs were isolated from citrate peripheral blood of vaccinated individuals by density gradient centrifugation using Biocoll^®^ separating solution, density 1.077 g/ml (Bio&Sell) and frozen in heat-inactivated FCS + 10% DMSO (Sigma-Aldrich) for liquid nitrogen storage. Thawed PBMCs were cultured in complete RPMI medium (RPMI 1640 medium (Thermo Fisher) supplemented with 10% heat-inactivated FCS, 1 mM ß-Mercaptoethanol, 1 mg/ml gentamicin, 23.83 g/l HEPES, 4.0 g/l L-glutamine, and 2000 U/ml penicillin-streptomycin) at 37°C and 5% CO_2_.

### IFN-γ Enzyme-linked immunospot (ELISPOT)

Cryopreserved PBMCs were thawed and rested overnight at 1×10^6^ cells/ml in complete RPMI medium. ELISPOT plates (Millipore) were coated with anti-human IFN-γ monoclonal antibody (clone 1-DIK, Mabtech) at 0.5 µg/well overnight at 4°C. Plates were washed with sterile PBS and subsequently blocked with complete RPMI medium for 1-2 h at 37°C. 400,000 PBMCs/well were seeded and stimulated with 11aa overlapping 15-mer PepMix™ SARS-CoV-2 spike glycoprotein peptide pool (1 µg/ml), provided in two peptide sub-pools S1 and S2 (JPT), for 20 h at 37°C. For the unstimulated condition, PBMCs were cultured in complete RPMI medium and respective dilution of solvent DMSO. As a positive control, PBMCs were stimulated with 25 ng/ml phorbol myristate acetate (PMA) (Sigma-Aldrich) and 1 µg/ml ionomycin (Sigma-Aldrich). Following this incubation, all steps were performed at room temperature. Plates were washed with PBS containing 0.05% Tween-20 (Sigma-Aldrich) and incubated with biotinylated anti-human IFN-γ monoclonal antibody (clone 7-B6-1, Mabtech) at 0.2 µg/well for 2 h. Following a second wash step with PBS containing 0.05% Tween-20, plates were incubated with an avidin-biotinylated peroxidase complex (VECTASTAIN^®^ Elite ABC-HRP Kit, Vector Laboratories) for 1-2 h. After final washing steps with first PBS containing 0.05% Tween-20 and then PBS, plates were developed by the addition of AEC substrate solution (Sigma-Aldrich) for 15 minutes. Subsequently, plates were washed with water, dried for 24 h in the dark, and analyzed on an ImmunoSpot^®^ Analyzer (Cellular Technologies Limited). A positive peptide-specific response was quantified by subtraction of mean spots of the unstimulated control and depicted as spot forming units (SFU)/10^6^ PBMCs.

### IFN-γ/IL-2 Fluorospot assay

Cryopreserved PBMCs were thawed and rested overnight at 2×10^6^ cells/ml in complete RPMI medium. Human IFN-γ/IL-2 Fluorospot assays (CTL Europe, Germany) were performed according to the manufacturer’s instructions. One day before the Fluorospot assays were performed, the plates were activated by adding 70% ethanol for less than one minute. Followed by a washing step and addition of IFN-γ/IL-2 capture antibodies overnight. After decanting the plate, 200,000 PBMCs/well were seeded and stimulated with 11aa overlapping 15-mer PepMix™SARS-CoV-2 spike glycoprotein peptide pool (1 µg/ml), provided in two peptide sub-pools S1 and S2 (JPT), for 20 h at 37°C. As antigen-specific positive control, we used a CEF pool of in total 32 15mer peptides derived from Cytomegalovirus (5 peptides), Epstein-Barr virus (15 peptides), and Influenza virus (Flu) (12 peptides) proteins (National Institute for Biological Standards and Control (NIBSC), UK). For the unstimulated condition, PBMCs were cultured in complete RPMI medium. After the stimulation period, the plates were washed and 80 µL of anti-human IFN-γ (FITC)/anti-human IL-2 (Hapten2) detection antibody solution was added for 2h at room temperature. For the visualization of secreted cytokines, plates were washed and a tertiary solution including anti-FITC Alexa Fluor® 488 (visualizes IFN-γ) and anti-Hapten2 CTL-Red™ (visualizes IL-2) was added for one hour. The staining procedure was stopped by washing the plate. After drying the plates for 24h on paper towels on bench top, Fluorospot plates were scanned and analyzed using an automated reader system (ImmunoSpot Ultimate UV Image analyzer/ImmunoSpot 7.0.17.0 Professional DC Software, CTL Europe GmbH, Germany). Positive reactivity to experimental stimulatory agents was given when the spot count in antigen-stimulated cells was greater than twice the spot count in unstimulated (background) wells.

### Intracellular cytokine staining (ICCS)

Cryopreserved PBMCs were thawed and rested overnight at 1×10^6^ cells/ml in complete RPMI medium. 10^6^ PBMCs were stimulated with spike glycoprotein peptide pool as described above for 20 h at 37°C in the presence of 1 µl/ml GolgiPlug™ (BD Biosciences). For the unstimulated condition, PBMCs were cultured in complete RPMI medium and respective dilution of solvent DMSO. As a positive control, PBMCs were stimulated with 25 ng/ml PMA and 1 µg/ml ionomycin. Following this incubation, all steps were performed at 4°C. PBMCs were washed twice with FACS buffer (PBS containing 0.05% BSA) and stained with ethidium-monoazide-bromide (EMA) (Thermo Fisher) for 15 minutes for live/dead discrimination. After two washing steps with FACS buffer, PBMCs were stained for surface markers CD8-eFluor450 (clone OKT8, Thermo Fisher, dilution 1:200) and CD4-PE (clone RPA-T4, Thermo Fisher, 1:400) for 20 minutes. Excess antibody was removed by two washing steps with FACS buffer followed by fixation/permeabilization using Cytofix/Cytoperm (BD Biosciences). PBMCs were washed twice with 1x Perm Wash buffer (BD Biosciences) and subsequently stained intracellularly for IL-2-APC (clone 5344.11, BD Biosciences, 1:20) and IFN-γ-FITC (clone 25723.11, BD Biosciences, 1:10) for 30 minutes. Following washing steps with first 1x Perm Wash buffer and then FACS-buffer, PBMCs were filtered through a nylon mesh and acquired on a LSRFortessa™ flow cytometer (BD Biosciences). A positive peptide-specific response was quantified by subtracting the mean frequency of IL-2 and IFN-γ double-positive T cells of the unstimulated control.

### Statistical analysis and graphical presentation

Statistics as well as figures were created with PRISM GraphPad 9.3.1.

## Data Availability

All data produced in the present study are available upon reasonable request to the authors.

## Author contributions

Conceptualization UP, OAC, KÜ, MT, PS

Methodology PI, PS, SB, JH, MW

Formal Analysis PI, MT, EV, AP, NK, CCC, AW, BHL, TB, JS,

Investigation PI, MT, EV, AP, NK, CCC, AW, BHL, SH, TB, RB, SJ, CC, SS, KT, SY,

PS

Resources KÜ, MT, UP, PK

Data Curation SB, PS, EV, MT, NK, CCC, AW, BHL, TB, HM, MW

Writing MT, KS, EV, KK, AP, UP, CB

Visualization AP, EV, KK, MT

Supervision KÜ, UP, MT, KS, NK, TB, PK; PS

Project Administration OAC, KÜ, UP, MT, CB

Funding Acquisition OAC, KÜ, UP

## Competing interests

JH reports grants and speaker honoraria from Pfizer, outside the study. UP reports grants from ALiOS and VirBio, and personal fees from AbbVie, Arbutus, Gilead, GSK, Johnson & Johnson, Roche, Sobi, and Vaccitech, outside the study. UP is co-founder and shareholder of SCG Cell Therapy OAC reports grants or contracts from Amplyx, Basilea, BMBF, Cidara, DZIF, EU-DG RTD (101037867), F2G, Gilead, Matinas, MedPace, MSD, Mundipharma, Octapharma, Pfizer, Scynexis; Consulting fees from Amplyx, Biocon, Biosys, Cidara, Da Volterra, Gilead, Matinas, MedPace, Menarini, Molecular Partners, MSG-ERC, Noxxon, Octapharma, PSI, Scynexis, Seres; Honoraria for lectures from Abbott, Al-Jazeera Pharmaceuticals, Astellas, Grupo Biotoscana/United Medical/Knight, Hikma, MedScape, MedUpdate, Merck/MSD, Mylan, Pfizer; Payment for expert testimony from Cidara; Participation on a Data Safety Monitoring Board or Advisory Board from Actelion, Allecra, Cidara, Entasis, IQVIA, Jannsen, MedPace, Paratek, PSI, Shionogi; A patent at the German Patent and Trade Mark Office (DE 10 2021 113 007.7); Other interests from DGHO, DGI, ECMM, ISHAM, MSG-ERC, Wiley, outside the submitted work.

All other authors declare no competing interests.

## Acknowledgments

The study was funded by the German Centre for Infection Research (DZIF), the European Union’s “Horizon 2020 Research and Innovation Programme” under grant agreement No. 101037867 (VACCELERATE), the “Bayerisches Staatsministerium für Wissenschaft und Kunst” for the CoVaKo-2021 and the For-COVID projects and the Helmholtz Association via the collaborative research program “CoViPa”. Further support was obtained from the Federal Ministry of Education and Science (BMBF) through the “Netzwerk Universitätsmedizin”, project “B-Fast” and “Cov-Immune”. KS is supported by the German Federal Ministry of Education and Research (BMBF, 01KI2013) and the Else-Kröner-Stiftung (2020_EKEA.127). We thank Manuela Stefanie Glocker, Thomas Wochnig, Romina Bester, Philip Hagen, Kirsten Fraedrich, Norbert Donhauser, Manuela Laumer for excellent technical assistance.

## Notes

### Author Declarations

The ethics committee of the medical faculty of the Friedrich-Alexander Universitat of Erlangen-Nurnberg, Germany, gave ethical approval for this work (Az. 340_21B). For the study side Munich, the ethic committee of the medical faculty of the Technische Universitat Munchen, Germany, gave ethical approval for tihs work (Az. 26/21 and Az. 330/21 S).

