## Supplemental information for "Dynamics of humoral and cellular immune responses after homologous and heterologous SARS-CoV-2 vaccination with ChAdOx1 nCoV-19 and BNT162b2"

**Supplemental Figure 1: Correlation between sVNT and rVNT using B.1.167.2** (A) Samples from the Erlangen study site collected at the late time point were used. The arbitrary units (AU)/ml of the sVNT were plotted against the IC50 values from the rVNT. The correlation was highly significant and reveals a spearman correlation coefficient of 0.7745 (95 % CI: 0.6632 – 0.8532) for all 78 data points.

**Supplemental Figure 2: Confirmation of serum neutralization capacity in a pseudovirus neutralization test.** 78 samples from the Erlangen study site collected at the late time point were analyzed by pVNT for the neutralization of either WT/D614G (A) or the Delta variant (B). Data points are shown for individual participants and bars represent group medians (ChAd/BNT n = 28, BNT-BNT n = 30, ChAd-ChAd n=20). The dashed line indicates the lower limit of detection. Data were analyzed by Kruskal–Wallis test (one-way ANOVA) followed by Dunn's multiple comparison. Statistically significant differences are indicated by p values (*p < 0.05; **p < 0.005; ***p < 0.0005; ****p < 0.0001).

**Supplemental Figure 3: Detection of serum antibodies able to bind SARS-CoV-2 Spike proteins of different variants.** Serum antibodies at the late time point for representative sub cohorts of participants, with n = 28 (ChAd-BNT), 30 (BNT-BNT) and 20 (ChAd-ChAd). S-specific IgG antibodies were determined against S-protein of the wildtype (WT), Delta or Omicron. The reference consists of a reconvalescent serum from the early phase of the pandemic which contains 1.01 mg/ml S (WT)-specific IgG. The median fluorescence intensity (MFI) is proportional to the amount of S-binding antibodies. Bars represent group medians overlaid with individual data points. Data were analyzed by Kruskal–Wallis test (one-way ANOVA) followed by Dunn's multiple comparison test. *p < 0.05, **p < 0.01, ***p < 0.001, ****p < 0.0001, and n.s. indicates not significant.

**Supplemental Figure 4: Intracellular cytokine staining (ICCS) for polyfunctional T cells after homologous and heterologous vaccination.** Flow cytometric analyses of polyfunctional spike-specific T cells after stimulation with SARS-CoV-2 spike peptide pools S1 and S2, in dilution of solvent (Neg. ctrl.), or with PMA/ionomycin (Pos. ctrl.). Vaccinees of Erlangen study center 4 months after secondary vaccination (late after #2). Representative flow cytometry data (left) and quantification of IL-2 and IFN-γ double-positive CD4 (A) and CD8 (B)T cells in technical duplicates (right). Shown gates are pre-gated for living CD4+ or CD8+ lymphocytes.

**Supplemental Figure 5: Correlation of SRAS-CoV-2 spike-specific T cells determined by ICCS and IFN-γ ELISPOT.** Correlation between IFN-γ spot-forming units (SFU) quantified by IFN-γ ELISPOT and IFN-γ+ T cells detected by intracellular cytokine staining. Each dot represents one vaccinee of the color-coded vaccination cohort. Correlation was measured by Spearman correlation. *$p < 0.05$, **$p < 0.01$, ***$p < 0.001$, ****$p < 0.0001$, and n.s. indicates not significant.

**Supplemental Figure 6: Long-term maintenance of polyfunctional T cells at prime vaccination level.** (A)-(D) Longitudinal characterization of polyfunctional spike-specific T cells, quantified by IFN-γ/IL-2 Fluorospot after stimulation with SARS-CoV-2 spike peptide pools S1 and S2. Vaccinees of Munich study center. (A) Representative data 4 months after second vaccination (late after #2) and longitudinal quantification of mono-functional IFN-γ secreting (B), mono-functional IL-2 secreting (C) and IFN-γ and IL-2 double-secreting (D) T cells. "early after #1": 55 to 137 days after first ChAd vaccination, n=26. "early after #2": 12 to 36 days after second BNT vaccination, n=29. "late after 2#": 91 to 153 days after second BNT vaccination, n=22. Dots represent individual vaccinees. Numbers indicate vaccinees with a positive response defined by a detectable T-cell response above background. Non-responsive vaccinees are represented as not detected (n.d.). Over time comparison within one group was done by Mann-Whitney test. *$p < 0.05$, **$p < 0.01$, ***$p < 0.001$, ****$p < 0.0001$, and n.s. indicates not significant.

**Supplement Figure 1**

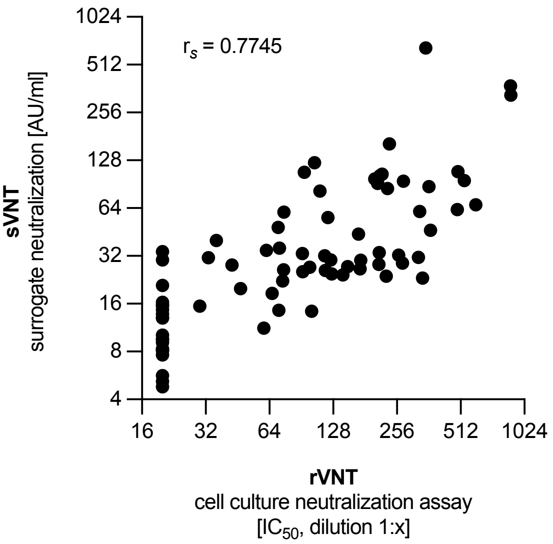

#### Supplement Figure 2

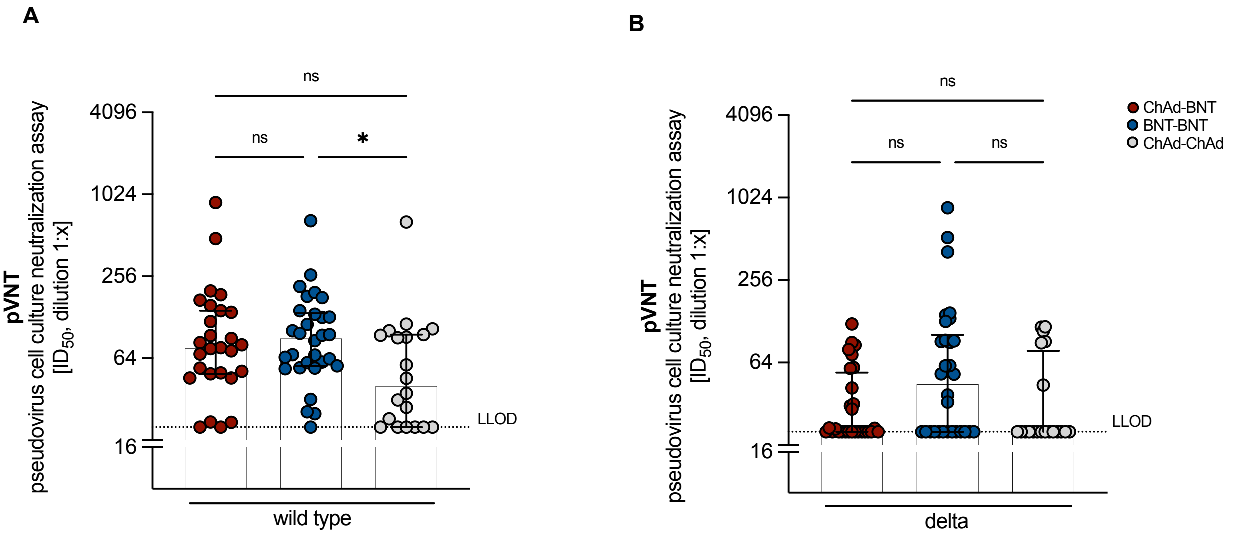

#### Supplement Figure 3

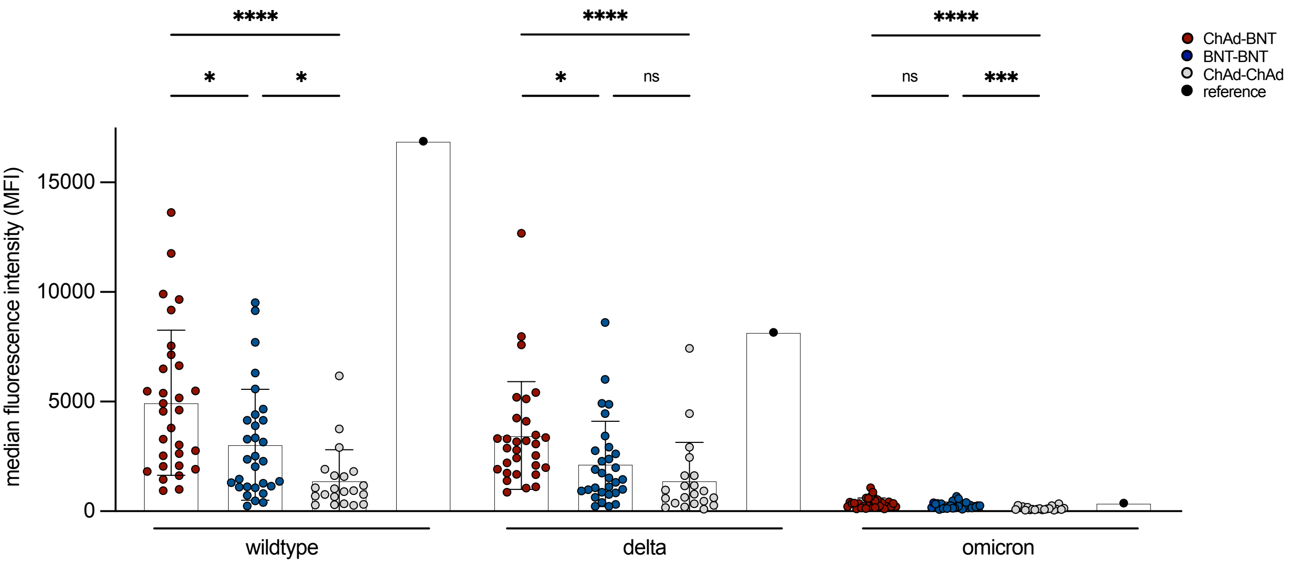

#### Supplement Figure 4

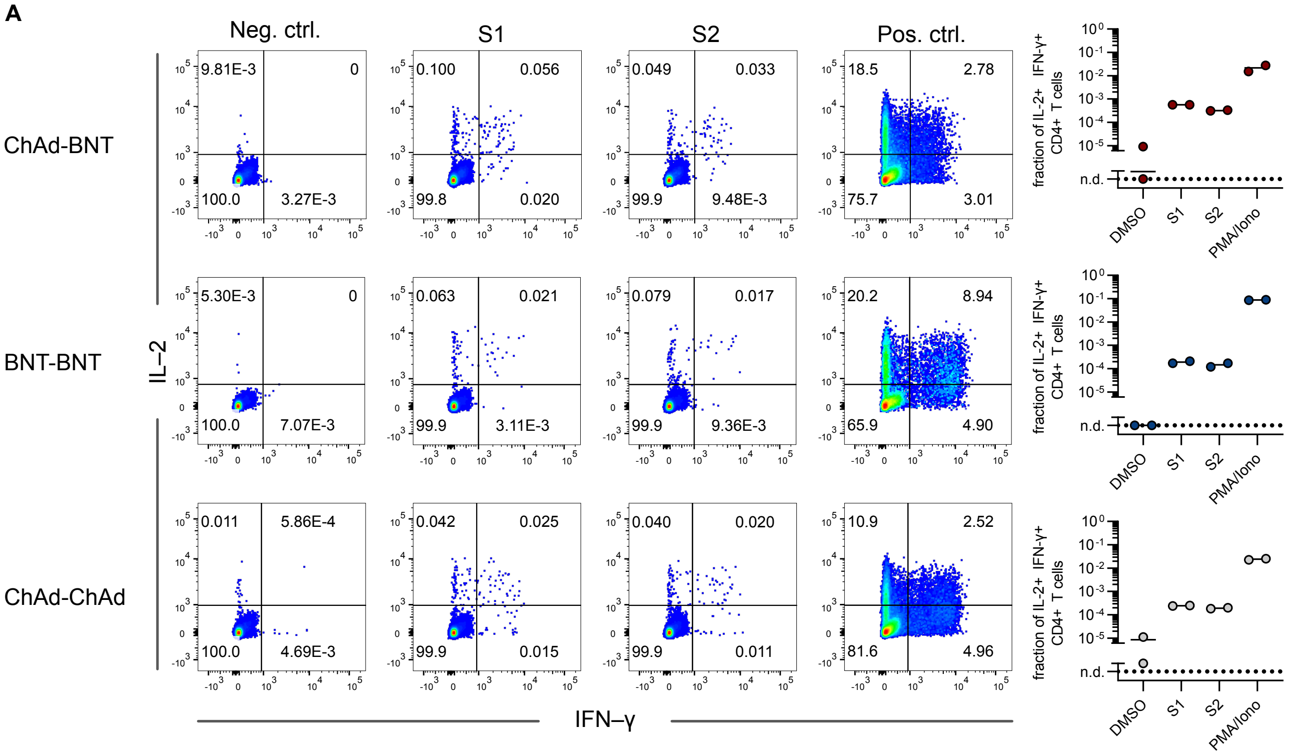

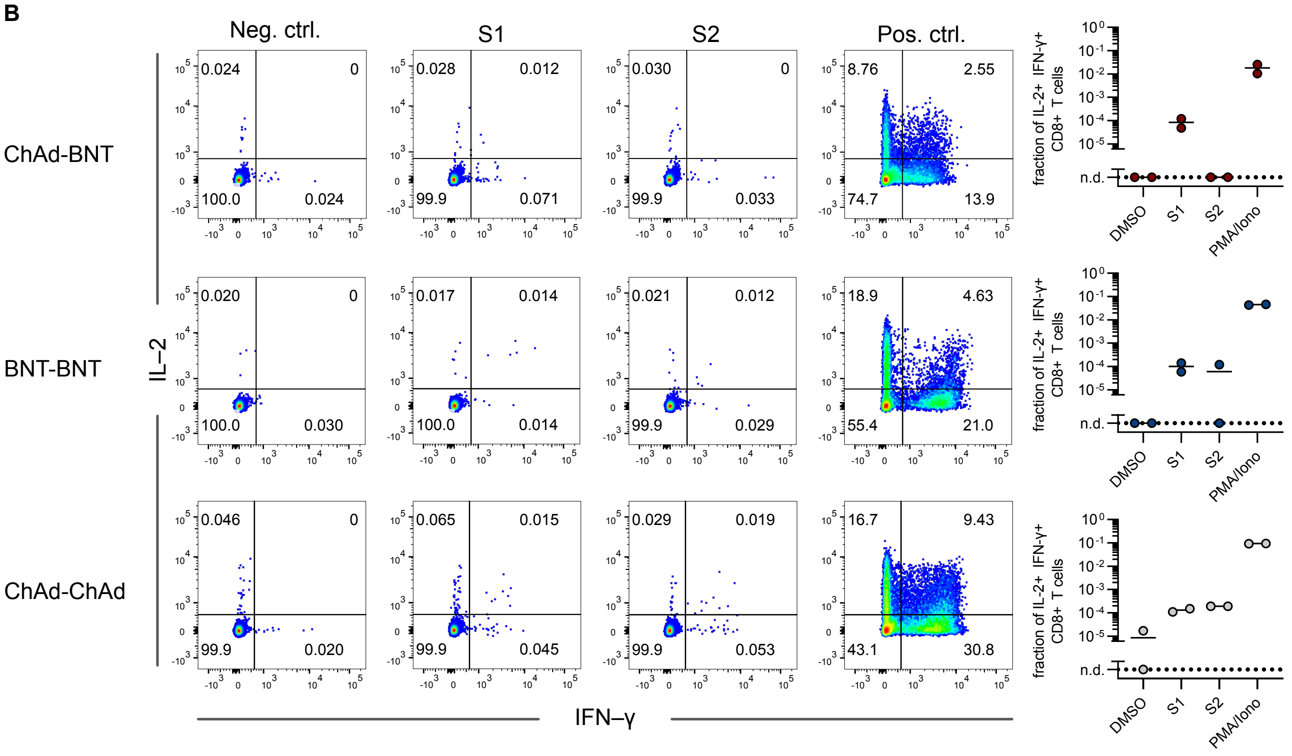

### Supplement Figure 5

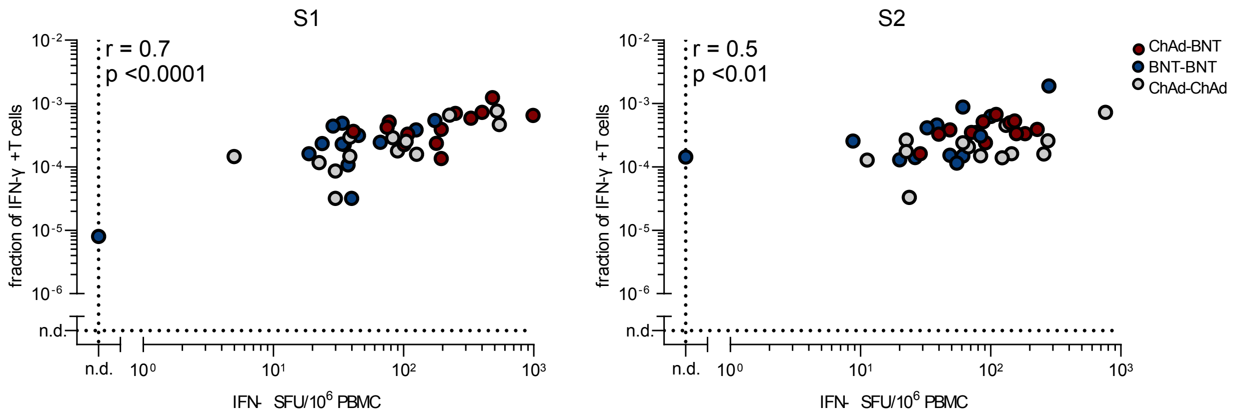

#### Supplement Figure 6

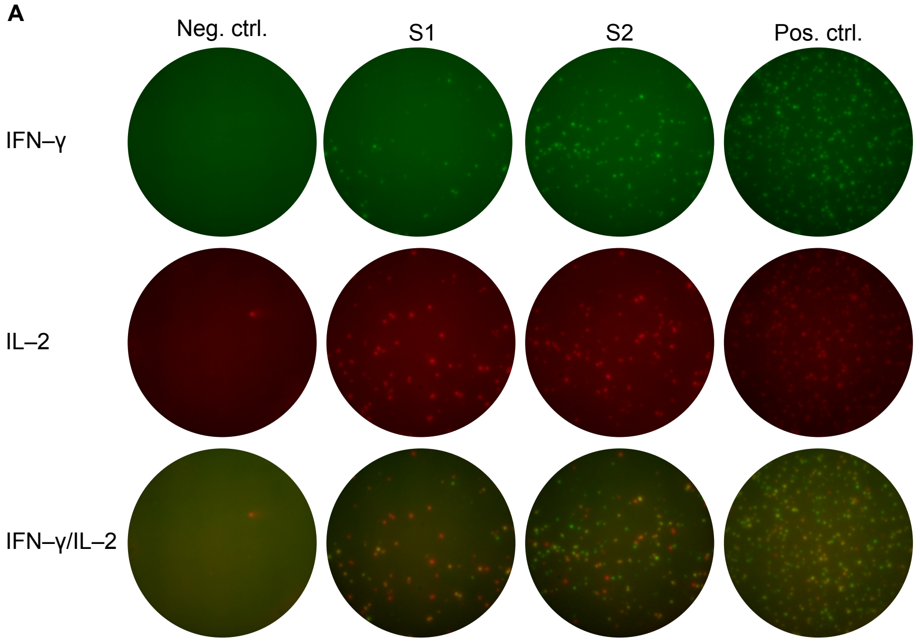

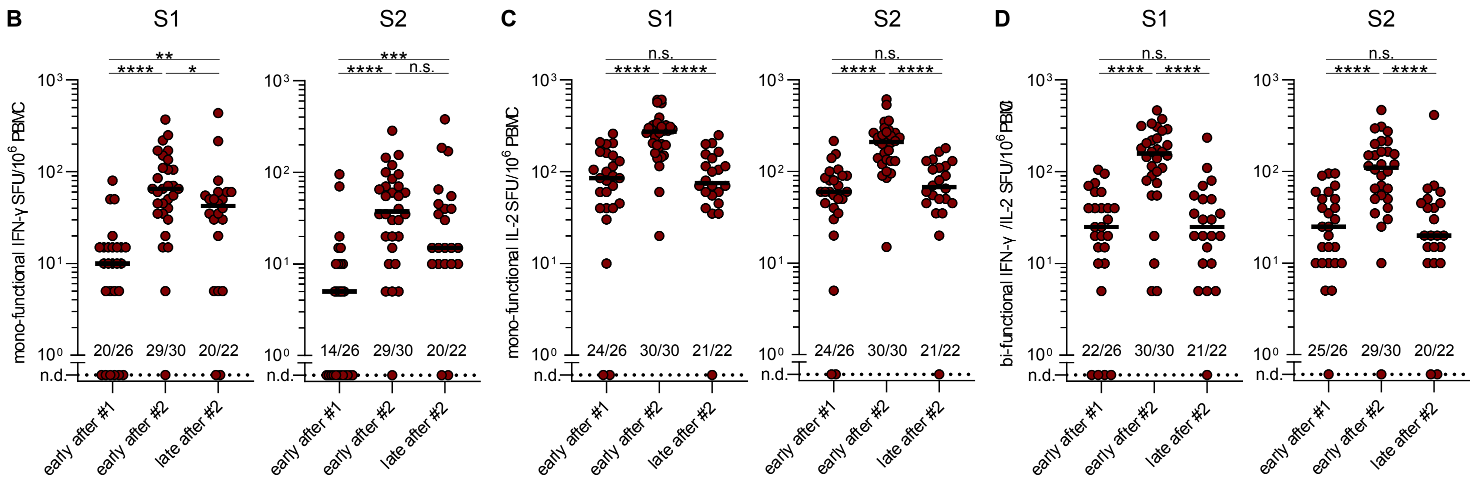

#### Supplemental Table for Figure 1A

| Supplemental Table for Figure 1A | | | | | | |
|---|---|---|---|---|---|---|
| | sVNT Median in AU/ml | IQR | N | Timespan Δ in days | Significance | |
| ChAd-BNT | 800 | 800 – 800 | 50 | 63 | | early after #2 |
| BNT-BNT | 791.31 | 708.58 - 800 | 50 | 21 | | |
| ChAd-BNT vs. BNT-BNT | | | | | ****P < 0.0001 | |
| ChAd-BNT | 234.65 | 160.93 - 500.83 | 43 | 110 | | late after #2 |
| BNT-BNT | 328.17 | 171.45 - 581.67 | 46 | 98 | | |
| ChAd-BNT vs. BNT-BNT | | | | | ns P = 0.3121 | |
| ChAd-$BNT_{early\#2}$ vs. ChAd-$BNT_{late\#2}$ | | | | | ****P < 0.0001 | over time |
| BNT-$BNT_{early\#2}$ vs. BNT-$BNT_{late\#2}$ | | | | | ****P < 0.0001 | |

#### Supplemental Table for Figure 1B

| Supplemental Table for Figure 1B | | | | | | |
|---|---|---|---|---|---|---|
| | sVNT Median in AU/ml | IQR | N | Timespan Δ in days | Significance | |
| ChAd-BNT | 762.78 | 484.46 - 800 | 201 | 63 | | early after #2 |
| BNT-BNT | 617.26 | 389.73 - 800 | 119 | 23 | | |
| ChAd-ChAd | 31.69 | 19.18 - 72.92 | 53 | 63 | | |
| ChAd-BNT vs. BNT-BNT | | | | | *P = 0.0381 | |
| ChAd-BNT vs. ChAd-ChAd | | | | | ****P < 0.0001 | |
| BNT-BNT vs. ChAd-ChAd | | | | | ****P < 0.0001 | |
| ChAd-BNT | 41.72 | 23.44 - 85.15 | 201 | 142 | | late after #2 |
| BNT-BNT | 31.74 | 21.05 - 74.38 | 119 | 158 | | |
| ChAd-ChAd | 9.32 | 6.01 - 14.38 | 53 | 142 | | |
| ChAd-BNT vs. BNT-BNT | | | | | ns P = 0.4989 | |
| ChAd-BNT vs. ChAd-ChAd | | | | | ****P < 0.0001 | |
| BNT-BNT vs. ChAd-ChAd | | | | | ****P < 0.0001 | |
| ChAd-$BNT_{early\#2}$ vs. ChAd-$BNT_{late\#2}$ | | | | | ****P < 0.0001 | over time |
| BNT-$BNT_{early\#2}$ vs. BNT-$BNT_{late\#2}$ | | | | | ****P < 0.0001 | |
| ChAd-$ChAd_{early\#2}$ vs. ChAd-$ChAd_{late\#2}$ | | | | | ****P < 0.0001 | |

#### Supplemental Table for Figure 2A

| **Supplemental Table for Figure 2A** | | | | | | | |
|---|---|---|---|---|---|---|---|
| | variant | rVNT Median in 1:x | IQR | N | timespan Δ in days | significance | |
| ChAd-BNT | B.1.617.2 | 929.15 | 419.7 - 2560 | 50 | 63 | | early after #2 |
| BNT-BNT | | 432.85 | 198.35 - 749.55 | 50 | 21 | | |
| ChAd-BNT vs. BNT-BNT | | | | | | ***P = 0.0003 | |
| ChAd-BNT | B.1.1.529 | 36 | 20 - 76.05 | 15 | 63 | | |
| BNT-BNT | | 20 | 20 - 20 | 15 | 21 | | |
| ChAd-BNT vs. BNT-BNT | | | | | | *P = 0.0122 | |
| ChAd-BNT | B.1.617.2 | 398.20 | 139.95 - 858.15 | 43 | 110 | | late after #2 |
| BNT-BNT | | 72.93 | 38.64 - 142.52 | 46 | 98 | | |
| ChAd-BNT vs. BNT-BNT | | | | | | ****P < 0.0001 | |
| ChAd-BNT | B.1.1.529 | 20 | 20 – 20 | 15 | 110 | | |
| BNT-BNT | | 20 | 0 - 20 | 15 | 98 | | |
| ChAd-BNT vs. BNT-BNT | | | | | | *P = 0.0105 | |
| ChAd-BNT$_{early\#2}$ vs. ChAd-BNT$_{late\#2}$ | B.1.617.2 | | | | | ***P = 0.0001 | over time |
| BNT-BNT$_{early\#2}$ vs. BNT-BNT$_{late\#2}$ | | | | | | ****P < 0.0001 | |
| ChAd-BNT$_{early\#2}$ vs. ChAd-BNT$_{late\#2}$ | B.1.1.529 | | | | | **P = 0.0039 | |
| BNT-BNT$_{early\#2}$ vs. BNT-BNT$_{late\#2}$ | | | | | | *P = 0.0156 | |

#### Supplemental Table for Figure 2B

| **Supplemental Table for Figure 2B** | | | | | | | |
|---|---|---|---|---|---|---|---|
| | variant | rVNT Median in 1:x | IQR | N | timespan Δ in days | significance | |
| ChAd-BNT | B.1.617.2 | 107.8 | 43.65 - 266.52 | 30 | 142 | | late after #2 |
| BNT-BNT | | 172 | 103.67 - 230.57 | 30 | 158 | | |
| ChAd-ChAd | | 20 | 20 – 70.8 | 21 | 142 | | |
| ChAd-BNT vs. BNT-BNT | | | | | | ns P = 0.8119 | |
| ChAd-BNT vs. ChAd-ChAd | | | | | | **P = 0.0072 | |
| BNT-BNT vs. ChAd-ChAd | | | | | | ***P = 0.0002 | |

#### Supplemental Table for Figure 3

| **Supplemental Table for Figure 3** | | | | | | |
|---|---|---|---|---|---|---|
| | Avidity in % | IQR | N | Timespan Δ in days | Significance | |
| ChAd-BNT | 57.96 | 47.03 - 62.55 | 12 | 63 | | early after #2 |
| BNT-BNT | 30.87 | 25.03 - 33.15 | 12 | 21 | | |
| ChAd-BNT vs. BNT-BNT | | | | | ****P < 0.0001 | |
| ChAd-BNT | 65.69 | 64.19 - 70.07 | 12 | 110 | | late after #2 |
| BNT-BNT | 49.49 | 42.91 - 55.97 | 12 | 98 | | |
| ChAd-BNT vs. BNT-BNT | | | | | ***P = 0.0004 | |
| ChAd-BNT$_{early\#2}$ vs. ChAd-BNT$_{late\#2}$ | | | | | ns P = 0.4118 | over time |
| BNT-BNT$_{early\#2}$ vs. BNT-BNT$_{late\#2}$ | | | | | ns P = 0.0537 | |

#### Supplemental Table for Supplemental Figure 1

| Supplemental Table for Supplemental Figure 1 | | | | |
|---|---|---|---|---|
| | Spearman r | 95 % CI | N | significance |
| rVNT vs. sVNT | 0.7745 | 0.6632 – 0.8532 | 78 | two tailed ****P < 0.0001 |

#### Supplemental Table for Supplemental Figure 2A and B

| Supplemental Table for Figure 2A and B | | | | | | |
|---|---|---|---|---|---|---|
| | variant | pVNT Median in 1:x | IQR | N | significance | |
| ChAd-BNT | wt | 76.55 | 49.4 – 143.6 | 27 | | early after #2 |
| BNT-BNT | | 90.62 | 56.06 – 137.1 | 30 | | |
| ChAd-ChAd | | 40.56 | 20 – 95.45 | 30 | | |
| ChAd-BNT vs. BNT-BNT | | | | | ns P > 0.9999 | |
| ChAd-BNT vs. ChAd-ChAd | | | | | ns P = 0.1872 | |
| BNT-BNT vs. ChAd-ChAd | | | | | *P = 0.0426 | |
| ChAd-BNT | B.1.617.2 | 20,50 | 20 – 54.03 | 28 | | |
| BNT-BNT | | 44.94 | 20 – 101.9 | 30 | | |
| ChAd-ChAd | | 20 | 20 – 78.03 | 20 | | |
| ChAd-BNT vs. BNT-BNT | | | | | ns P = 0.3165 | |
| ChAd-BNT vs. ChAd-ChAd | | | | | ns P > 0.9999 | |
| BNT-BNT vs. ChAd-ChAd | | | | | ns P = 0.1512 | |

#### Supplemental Table for Supplemental Figure 3

| Supplemental Table for Supplemental Figure 3 | | | | | | |
|---|---|---|---|---|---|---|
| | variant | MFI | IQR | N | significance | |
| ChAd-BNT | wt | 4586 | 2072 – 6767 | 30 | | early after #2 |
| BNT-BNT | | 2328 | 1111 – 4218 | 30 | | |
| ChAd-ChAd | | 941.8 | 509 – 1721 | 21 | | |
| ChAd-BNT vs. BNT-BNT | | | | | *P = 0.0438 | |
| ChAd-BNT vs. ChAd-ChAd | | | | | ****P < 0.0001 | |
| BNT-BNT vs. ChAd-ChAd | | | | | *P = 0.0205 | |
| ChAd-BNT | B.1.617.2 | 2972 | 1874 – 4138 | 30 | | |
| BNT-BNT | | 1489 | 861.8 – 2799 | 30 | | |
| ChAd-ChAd | | 785.6 | 358.8 – 1624 | 21 | | |
| ChAd-BNT vs. BNT-BNT | | | | | *P = 0.0162 | |
| ChAd-BNT vs. ChAd-ChAd | | | | | ****P < 0.0001 | |
| BNT-BNT vs. ChAd-ChAd | | | | | ns P = 0.1659 | |
| ChAd-BNT | B.1.1.529 | 303 | 212.9 – 488.7 | 30 | | |
| BNT-BNT | | 228.1 | 154.1 – 365.6 | 30 | | |
| ChAd-ChAd | | 99.88 | 73.87 – 183.5 | 21 | | |
| ChAd-BNT vs. BNT-BNT | | | | | ns P = 0.4610 | |
| ChAd-BNT vs. ChAd-ChAd | | | | | ****P < 0.0001 | |
| BNT-BNT vs. ChAd-ChAd | | | | | ***P = 0.0009 | |
